# Mycotoxin exposure biomonitoring in breastfed and non-exclusively breastfed Nigerian children

**DOI:** 10.1101/2020.05.28.20115055

**Authors:** Chibundu N. Ezekiel, Wilfred A. Abia, Dominik Braun, Bojan Šarkanj, Kolawole I. Ayeni, Oluwawapelumi A. Oyedele, Emmanuel C. Michael-Chikezie, Victoria C. Ezekiel, Beatrice N. Mark, Chinonso P. Ahuchaogu, Rudolf Krska, Michael Sulyok, Paul C. Turner, Benedikt Warth

**Author notes:** Corresponding authors: Chibundu N. Ezekiel, Department of Microbiology, Babcock University, Ilishan Remo, Ogun State, Nigeria., Benedikt Warth, University of Vienna, Faculty of Chemistry, Department of Food Chemistry and Toxicology, Währinger Straße 38, 1090 Vienna, Austria.

## Abstract

A multi-specimen, multi-mycotoxin approach involving ultra-sensitive LC-MS/MS analysis of breast milk, complementary food and urine was applied to examine mycotoxin co-exposure in 65 infants, aged 1–18 months, in Ogun state, Nigeria. Aflatoxin M_1_ was detected in breast milk (4/22 (18%)), while six other classes of mycotoxins were quantified; including dihydrocitrinone (6/22 (27%); range: 14.0–59.7ng/L) and sterigmatocystin (1/22 (5%); 1.2ng/L) detected for the first time. Seven distinct classes of mycotoxins including aflatoxins (9/42 (21%); range: 1.0– 16.2µg/kg) and fumonisins (12/42 (29%); range: 7.9–194µg/kg) contaminated complementary food. Mycotoxins covering seven distinct classes with diverse structures and modes of action were detected in 64/65 (99%) of the urine samples, demonstrating ubiquitous exposure. Two aflatoxin metabolites (AFM_1_ and AFQ_1_) and FB_1_ were detected in 6/65 (9%), 44/65 (68%) and 17/65 (26%) urine samples, respectively. Mixtures of mycotoxin classes were common, including 22/22 (100%), 14/42 (33%) and 56/65 (86%) samples having 2–6, 2–4, or 2–6 mycotoxins present, for breast milk, complementary food and urine, respectively. Aflatoxin and/or fumonisin was detected in 4/22 (18%), 12/42 (29%) and 46/65 (71%) for breast milk, complimentary foods and urine, respectively. Furthermore, the detection frequency, mean concentrations and occurrence of mixtures were typically greater in urine of non-exclusively breastfed compared to exclusively breastfed infants. The study provides novel insights into mycotoxin co-exposures in early-life. Albeit a small sample set, it highlights transition to higher levels of infant mycotoxin exposure as complementary foods are introduced, providing impetus to mitigate during this critical early-life period and encourage breastfeeding.

## 1. Introduction

Nutrition within the first 1000 days of life is crucial to the growth, development and performance of children in their later years and into adulthood as postulated in the ‘developmental origins of health and disease’ (DOHaD) concept (Barker, 2000; Bateson et al., 2004; Mandy and Nyirenda, 2018). However, the sources of children’s diets within this period may be diverse, and depend on the age of the children. The dietary patterns in a progressive manner include exclusively consumed breast milk or formula milk, breast milk or formula milk combined with complementary foods, and exclusively consumed complementary foods. Breast milk composition varies widely depending on diet, age, lactation stage, number of pregnancies, and other physiological parameters (Butts et al., 2018; Innis, 2014; Wu et al., 2018). Previous studies have shown that complementary foods can be made from single or mixed cereals as a porridge, sometimes in combination with nuts, animal milk and other mashed items, and additional food groups are added as children get older (Alvito et al., 2010; Chuisseu Njamen et al., 2018; Kamala et al. 2016; Kimanya et al., 2009, 2010, 2014; Ojuri et al., 2018, 2019). Unfortunately, in some parts of the world, some commonly consumed foods contain toxic fungal metabolites known as mycotoxins (Alvito et al., 2010; Braun et al., 2018; Cherkani-Hassani et al., 2016; Juan et al., 2014; Kamala et al., 2016; Kimanya et al., 2009, 2010, 2014; Chuisseu Njamen et al., 2018; Ojuri et al., 2018, 2019; Preindl et al., 2019; Warth et al., 2016).

Aflatoxin (AF), fumonisin (FUM), ochratoxin A (OTA), deoxynivalenol (DON) and zearalenone (ZEN) are considered priority mycotoxins and are regulated in food. AFs are categorized as class 1 human liver carcinogen (IARC, 2002, 2012a). AF is additionally implicated in growth faltering of infants, (Gong et al., 2002, 2003, 2004, 2012; Turner et al., 2003, 2007; Watson et al., 2018), possibly associated with interference to micronutrient absorption, impairment of gut integrity, liver homeostasis or effects on the IGF-growth axis (IARC, 2015; Turner, 2013; Watson et al., 2018). FUM intake has been associated with the incidence of esophageal cancer (Chu et al., 1994; Sun et al., 2007, 2011; Yoshizawa et al., 1994), birth defects (Missmer et al., 2006), and suggested to play a role in child growth faltering (Shirima et al., 2015). OTA is linked to renal toxicity (Heussner and Bingle, 2015), while immune system modulation was reported for trichothecenes such as DON (Pestka, 2010). Furthermore, *in vitro* data suggest interactions of regulated mycotoxins with emerging ones such as alternariol (AOH) and its methylated ether form (alternariol monomethyl ether; AME), beauvericin (BEA), enniatins and moniliformin (MON) (Aichinger et al., 2019; Vejdovszky et al., 2017a, 2017b; Woelflingseder, 2019).

Complementary foods can be contaminated with mycotoxins (Cappozzo et al., 2017; Chuisseu Djamen et al., 2018; Juan et al., 2014; Kimanya et al., 2009, 2010, 2014; Kolakowski et al., 2016; Ojuri et al., 2018, 2019; Oueslati et al., 2018), and ingested mycotoxins or their metabolites may be found in various bio-fluids including blood, urine and breast milk (Ayelign et al., 2017; Braun et al., 2018; Chen et al., 2017; Cherkani-Hassani et al., 2016; Ediage et al., 2013; Ezekiel et al., 2014; Ferrufino-Guardia et al., 2019; Gerding et al., 2015; Heyndrickx et al., 2015; Papageorgiou et al., 2018; Polychronaki et al., 2008; Sanchez and Diaz, 2019; Schwartzbord et al., 2016; Shirima et al., 2015; Turner et al., 2012a; Warth et al., 2016). Only a limited number of studies have assessed mycotoxin exposure in Nigerian children using biomarkers of exposure (Adejumo et al., 2013; Braun et al., 2018; Ezekiel et al., 2014, 2018b; McMillan et al., 2018; Sarkanj et al., 2018)-.

This study aimed at elucidating mycotoxin co-exposure patterns in infants using three measures (food, breast-milk and urine), all with comprehensive quantitative and ultra-sensitive mycotoxin analysis, to compare exposure levels in exclusively breastfed (EB) with non-exclusively breastfed (NEB) infants. Importantly, several classes of mycotoxins were examined, for example aflatoxins, and in bio-fluids (milk or urine) metabolites of such mycotoxins may also be observed in addition or instead of the parent class of toxin, for example aflatoxin M1 or Q1. These metabolites would not be observed in the original food.

## 2. Materials and methods

### 2.1 Study area and population

The study population was 65 infants aged 1–18 months from Ilishan and Ikenne in Remo land of Ogun state (south-western Nigeria). Both are small sister communities within 1 km distance of each other, with rich diversity of foods available in households. Both communities are semi-urban due to the presence of campuses of tertiary institutions and they share similar characteristics as detailed below. Indigent families in these communities are farmers and traders, and both communities consist of approximately 15,000 residents (Ezekiel et al., 2018b). The food sources for families are local markets, small stores and own farms. Major foods consumed are cereals (rice, maize, wheat), tubers (cassava and yam), and peanuts, melon seed and cowpea. The dietary patterns for infants in both communities involve exclusive breastfeeding (feeding infants with only breast milk for at least six months from birth) and non-exclusive breastfeeding (the introduction of complementary foods to breastfed infants sometimes from the first month after birth). Dietary pattern depends on family income, mother’s health and work status, and knowledge on breastfeeding practices. A majority of the families associated with tertiary institutions in both communities practice exclusive breastfeeding.

### 2.2 Sampling design and ethical considerations

This pilot, cross-sectional survey was conducted between January and February 2016. Families that participated were identified through local health centres and post-natal clinics in the two communities where mother/infant pairs were registered. The study consisted of two cohorts: exclusively breastfed (EB) children (age: 1–6 months) and non-exclusively breastfed (NEB) children (age: 3–18 months) to which 23 and 42 children (male/female ratio = 31:34) were recruited, respectively. A total of 50 and 15 infants were recruited into the study from Ilishan and Ikenne communities, respectively. The inclusion criteria for this study were age of infants and dietary patterns for each cohort as well as healthy status of infants. The age of infants recruited into the study were ascertained from their health records. Dietary patterns (exclusively breast milk consumption in EB cohort *versus* consumption of breast milk and complementary food in NEB cohort) were ascertained from the mothers. Infants with known medical conditions (e.g., jaundice, HIV positive or visibly malnourished) were excluded during the recruitment stage as it was the purpose of this study to investigate typical background exposure levels and compare EB with NEB rather than targeting the impact of exposure on health status. Prior to recruitment or inclusion into the study, the mother of each child was informed in her preferred language (English or Yoruba) on the purpose of the study. Only mothers who signed the informed consent document (and on behalf of their infant) to participate in the research were included into the study.

Complementary foods were obtained as “plate-ready” samples (*n*=42; 50 g each), by a trained team member, for the NEB children (*n*=42) on the day prior to infant urine sample collection. The food samples collected included industrially-processed infant cereal (*n*=6), fermented maize gruel *ogi* (*n*=26) and *tombran* (home-made pudding from mixed cereal and nut; *n*=10). One food sample representing the most frequently consumed food in a day was collected per infant. Breast milk samples (*n*=22; 3–5 mL each), considered as food for the EB infants, were collected from mothers of EB infants on the day preceding the sampling of infant urine. Complementary food and breast milk sample collections on the day prior to infant urine sampling were necessary in order to ensure adequate metabolism and transfer of toxins into urine. One mother did not provide breast milk sample due to intake of medications. Other details of sampling and handling of the breast milk samples were described by Braun et al. (2018). In this paper, the breast milk samples have been previously analysed for mycotoxins utilizing a less sensitive method. In addition, some of the data from the mother-infant pairs with exclusively breastfed infants were used in a recent correlation study by Braun et al. (*in revision*) that added longitudinal aspects and urinary exposure measurements of the mothers.

First morning urine samples (10–40 mL each) were collected in 50 falcon tubes from all 65 infants recruited into the study by the mothers, using their experience and knowledge of the time at which their infant pass urine in the morning. All samples (plate-ready complementary food, breast milk and urine) were immediately frozen at –20 °C and sent by courier on dry ice to Austria for LC-MS/MS analyses.

Ethical permission for this study was granted by the Babcock University Health Research Ethics Committee under two approval numbers: BUHREC294/16 for the EB children and BUHREC156/15 for the NEB children. In adherence to ethical standards, all samples were blinded and coded to exclude participant information prior to analytical measurements.

### 2.3 Demographic, dietary and health status questionnaire

Each mother received a copy of a well-structured questionnaire for completion prior to food, breast milk and urine sample collection. Copies of the questionnaire were administered by trained interviewers and translation to the local language (Yoruba) was available on request of some mothers. The administered questionnaire was designed to elicit information on participant demography, family socio-economic status, individual dietary preferences, food consumption frequencies and quantities, and associated health implications among the infants.

### 2.4 Mycotoxin determination in breast milk samples

Mycotoxins were quantified in breast milk samples as described in Braun et al. (2020). Briefly, each homogenized breast milk sample (1 mL) was extracted using 1 mL acidified ACN (1% formic acid) on a shaker for 3 min, followed by addition of anhydrous magnesium sulfate (0.4 g) and sodium chloride (0.1 g). Sequel to two rounds of cold centrifugation (4,750 x g at 10 °C for 10 min; 14,000 x g, 4 °C for 2 min), a SPE clean up step using SPE column (Oasis PRiME HLB^®^, Waters, Milford, MA, USA) followed. Mycotoxins were then concentrated on a vacuum concentrator (Labconco, Missouri, USA) prior to reconstitution using 81 µL MeOH/CAN and 9 µL of the internal standard mixture. Mycotoxins in breast milk samples were measured on a Sciex QTrap 6500^+^ LC-MS/MS system (Foster City, CA, USA) equipped with a Turbo-V™ electrospray ionization (ESI) source coupled to an Agilent 1290 series UHPLC system (Waldbronn, Germany). Analytes were separated on an Acquity HSS T3 column (2.1×100mm; Waters, Vienna, Austria) with 1.8 µm particle size at 40 °C. ESI-MS/MS was performed in scheduled multiple reaction monitoring (MRM) mode using fast polarity switching. For each analyte two individual mass transitions were acquired.

The analytical method was in-house validated according to the European Commission Decision 2002/657/EC (EC 2002) and the EuraChem Laboratory Guide (Magnusson, 2014). Limits of detection (LOD) and quantification (LOQ) are reported in Table 2 and the relative standard deviation as obtained during validation was below 9% for all mycotoxins when spiked approximately at their respective LOQ value (Braun et al., 2020). Quality control measures in routine analysis include a pooled non-spiked breast milk sample and three breast milk samples, which are fortified with authentic reference standards at the same concentration levels as during method validation to ensure proper extraction efficiency and instrumental performance.

### 2.5 LC-MS/MS-based mycotoxin analysis of food samples

Plate-ready complementary food samples were analysed for the presence of multiple mycotoxins according to the dilute and shoot LC-MS/MS-based method described by Sulyok et al. (2020). This method covers the detection of more than 500 metabolites, inclusive of mycotoxins, in food samples. Briefly, 5 g of each food sample was extracted with 20 mL of acetonitrile/water/acetic acid 79:20:1, (v/v/v) in a 50 mL polypropylene tube (Sarstedt, Nümbrecht, Germany) for 90 min on a GFL 3017 rotary shaker (GFL, Burgwedel, Germany). Afterwards, the extract was diluted 1:1 (v/v) in dilution solvent (acetonitrile/water/acetic acid 20:79:1, v/v/v) prior to measurement on a Sciex QTrap 5500 LC-MS/MS System (Applied Biosystem, Foster City, CA, USA) equipped with TurboIonSpray electrospray ionisation (ESI) source and a 1290 Series HPLC System (Agilent, Waldbronn, Germany). Chromatographic separation was performed at 25°C on a Gemini® C18-column, 150×4.6 mm i.d., 5 μm particle size, equipped with a C18 4×3 mm i.d. security guard cartridge (Phenomenex, Torrance, CA, USA). ESI-MS/MS was performed in the scheduled MRM mode both in positive and negative polarities in two separate chromatographic runs per sample by scanning two fragmentation reactions per analyte. The MRM detection window of each analyte was set to its expected retention time ±20 and ±26 s in the positive and the negative modes, respectively. The identification of each positive analyte was confirmed when two MRMs per analyte was obtained (EC, 2002). Further details on the method performance are described in Sulyok et al. (2020).

The analytical method has been fully validated following the SANTE Guide (SANTE, 2018) for multi-residue analysis and published (Sulyok et al., 2020). The latter reference is available open access and provides full information on all data on method performance. The ongoing quality control of the method involves participation in a proficiency testing scheme organized by BIPEA. Furthermore, a quality control check sample is included in each analytical sequence. The extended measurement uncertainty has been determined to be 50% (Stadler et al., 2018).

### 2.6 Urinary mycotoxin measurements by UPLC-MS-MS

Mycotoxins in the urine samples were determined following a stable-isotope dilution assay-based LC-MS/MS method described in detail by Šarkanj et al. (2018). Briefly, each urine sample was centrifuged for 3 min at 5600 x *g*, then treated β-glucuronidase from *E. coli* Type IX-A (Sigma-Aldrich, G7396-2MU) of prior to a SPE cleanup on Oasis PRiMEHLB® SPE columns (Waters, Milford, MA, USA). Extracts were then evaporated under nitrogen, reconstituted with 470 µL dilution solvent (10% acetonitrile, 0.1% glacial acetic acid) and fortified with 30 µL of an IS mixture. Measurement of mycotoxins in urine samples was performed on a Sciex QTrap 6500^+^ LC-MS/MS system (Foster City, CA) equipped with a Turbo-V™ ESI source coupled to an Agilent 1290 series UHPLC system (Waldbronn, Germany). Analytes were separated on an AtlantisT3 HSS column (2.1×100mm; Waters, Wexford, Ireland) with 1.8 µm particle size at 35 °C. ESI-MS/MS was performed in scheduled MRM mode and with a 180 sec detection window. At least two individual transitions were monitored for each analyte.

The analytical method was in-house validated according to the European Commission Decision 2002/657/EC (EC, 2002). LOD and LOQ values are presented in Table 4. Inter-day relative standard deviation during method validation was calculated to be below 33% for all mycotoxins included in this assay (Šarkanj et al., 2018). During routine analysis, QC samples were assessed on a regularly basis to ensure proper sample preparation and instrumental performance. After each 20 experimental samples a pooled fortified and non-fortified (‘blank’) urine sample were measured.

### 2.7 Data analysis

The IBM SPSS Statistics v21.0 (SPSS, Inc., Chicago, IL, USA) was applied in the analyses of all data. A scatter plot was created for the percentage increases in frequencies and means of urinary mycotoxins between the two cohorts of children. For comparison of urinary mycotoxin levels in the population, means were separated by the Duncan’s Multiple Range test (DMRT) and tested for significance by analysis of variance (ANOVA) at α = 0.05. For the comparison of all other data between the two cohorts of children in the study, the independent sample *t*-test using the Levene’s test for equality of variances were applied to test for significance at α = 0.05.

## 3. Results

### 3.1 Demography and food preference of children

The demographic characteristics of the infants as well as their food consumption data are given in Table 1. The NEB infants were significantly (*p*<0.01) older (9.0±3.6 months) and heavier (8.0±1.6 Kg; *p*<0.05) than the EB children (3.7±1.6 months and 6.3±1.0 Kg, respectively). All infants received breast milk at least once daily, with higher consumption frequency of 8–10 times per day in the EB cohort compared to 1–3 times per day in the NEB cohort who received complementary food at least thrice per day. NEB infants received a variety of foods, which in most cases (>90%) included infant cereal (mix of animal milk with maize, rice, or wheat), *ogi* (fermented maize gruel) and *tombran* (mixed cereal and nut pudding).

**Table 1.** Demographic characteristics and food preference of study participants.

| Characteristics | All | Exclusively<br>breastfed | Non-exclusively<br>breastfed |
| --- | --- | --- | --- |
| Subjects (*n* (%)) | 65 | 23 (35.4) | 42 (64.6) |
| Gender (*n* (%)) |  |  |  |
| Male | 31 (47.7) | 11 (47.8) | 20 (47.6) |
| Female | 34 (52.3) | 12 (52.2) | 22 (52.4) |
| Age (months) |  |  |  |
| Mean ± SD<sup>a</sup> | 7.1 ± 3.9 | 3.7 ± 1.6 | 9.0 ± 3.6* |
| Range | 1–18 | 1–6 | 3–18 |
| Body weight (kg) |  |  |  |
| Mean ± SD<sup>a</sup> | 7.4±1.7 | 6.3±1.0 | 8.0±1.6** |
| Range | 4.5–11.2 | 4.5–8.0 | 5.0–11.2 |
| Frequency of food consumption/day |  |  |  |
| Breast milk | 1–>10 | 8–>10<sup>b</sup> | 1–3 |
| Complementary food | 3–5 | 0 | 3–5 |
| Complementary food (*n* (%)) preference |  |  |  |
| Single food (e.g. *ogi*) | 3 (4.6) | 0 (0) | 3 (7.1) |
| Mixed foods<sup>c</sup> | 39 (60.0) | 0 (0) | 39 (92.9) |

<sup>a</sup>Standard deviation.
<sup>b</sup>As reported by Braun et al. (2018).
<sup>c</sup>Combination of complementary foods (e.g. infant cereal, *ogi* and *Tombran*) consumed in a day.
\*Significant at $p < 0.01$ , \*\* $p < 0.05$

**Table 2.** Mycotoxins in breast milk (*n* = 22) consumed by exclusively breastfed infants in Ogun state, Nigeria.

| Mycotoxins | LOD<br>(ng/L) | LOQ<br>(ng/L) | $N$ (%)<sup>a</sup> | Concentration (ng/L) | | |
| --- | --- | --- | --- | --- | --- | --- |
|  |  |  |  | Range | Median | Mean |
| Aflatoxin M<sub>1</sub> | 2.0 | 4.0 | 4 (18.2) | 2.0<sup>b</sup> | 2.0 | 2.0 |
| Alternariol monomethyl ether | 0.5 | 1.0 | 21 (95.5) | 0.5<sup>b</sup>–11.7 | 1.8 | 3.0 |
| Beauvericin | 0.1 | 0.3 | 22 (100) | 1.0–12.0 | 2.2 | 3.0 |
| Dihydrocitrinone | 14.0 | 28.0 | 6 (27.3) | 14.0<sup>b</sup>–59.7 | 14.0 | 25.2 |
| Enniatin B | 0.7 | 1.4 | 16 (72.7) | 0.7<sup>b</sup>–10.1 | 4.6 | 5.0 |
| Enniatin B<sub>1</sub> | 0.5 | 1.0 | 5 (22.7) | 0.5<sup>b</sup>–1.15 | 0.5 | 0.6 |
| Ochratoxin A | 2.0 | 4.0 | 14 (63.6) | 2.0<sup>b</sup>–67.6 | 2.0 | 9.6 |
| Ochratoxin B | 2.5 | 5.0 | 2 (9.1) | 5.3–6.7 | 6.0 | 6.0 |
| Sterigmatocystin | 0.5 | 1.0 | 1 (4.6) | 1.2 | 1.2 | 1.2 |

<sup>a</sup>Number (and percentage) of samples containing the mycotoxin.
<sup>b</sup><LOQ values were considered as positive data and substituted with LOQ/2.

### 3.2 Occurrence of mycotoxins in breast milk

Breast milk samples were examined for the presence of 34 mycotoxins (or their metabolites), of which nine were observed. All 22 breast milk samples contained mycotoxins although mostly at very low concentrations (Table 2). The most frequently detected mycotoxins were BEA (incidence: 100%; range: 1.0–12.0 ng/L; mean: 3.0 ng/L) and AME (incidence: 96%; range: 0.5–11.7 ng/L; mean: 3.0 ng/L), whereas DHC (incidence: 27%; range: 14.0–59.7 ng/L; mean: 25.2 ng/L) and OTA (incidence: 64%; range: 2.0–67.6 ng/L; mean: 9.6 ng/L) had higher mean levels than all other mycotoxins found in the samples. AFM_1_ was the only aflatoxin metabolite detected in breast milk, occurring in 18% of the samples at levels below the LOQ; thus, the data points were assigned LOQ/2 values of 2.0 ng/L (Table 2). Enniatins (EnnB and EnnB_1_) occurred in 77% of the samples, with EnnB dominating in 73% of samples at mean concentration of 5.0 ng/L (range: 0.7–10.1 ng/L). Sterigmatocystin (STER) was detected in one sample at 1.2 ng/L. The chromatograms of DHC and STER detected in breast milk are shown in Figure 1 (C–D).

**Figure 1.**
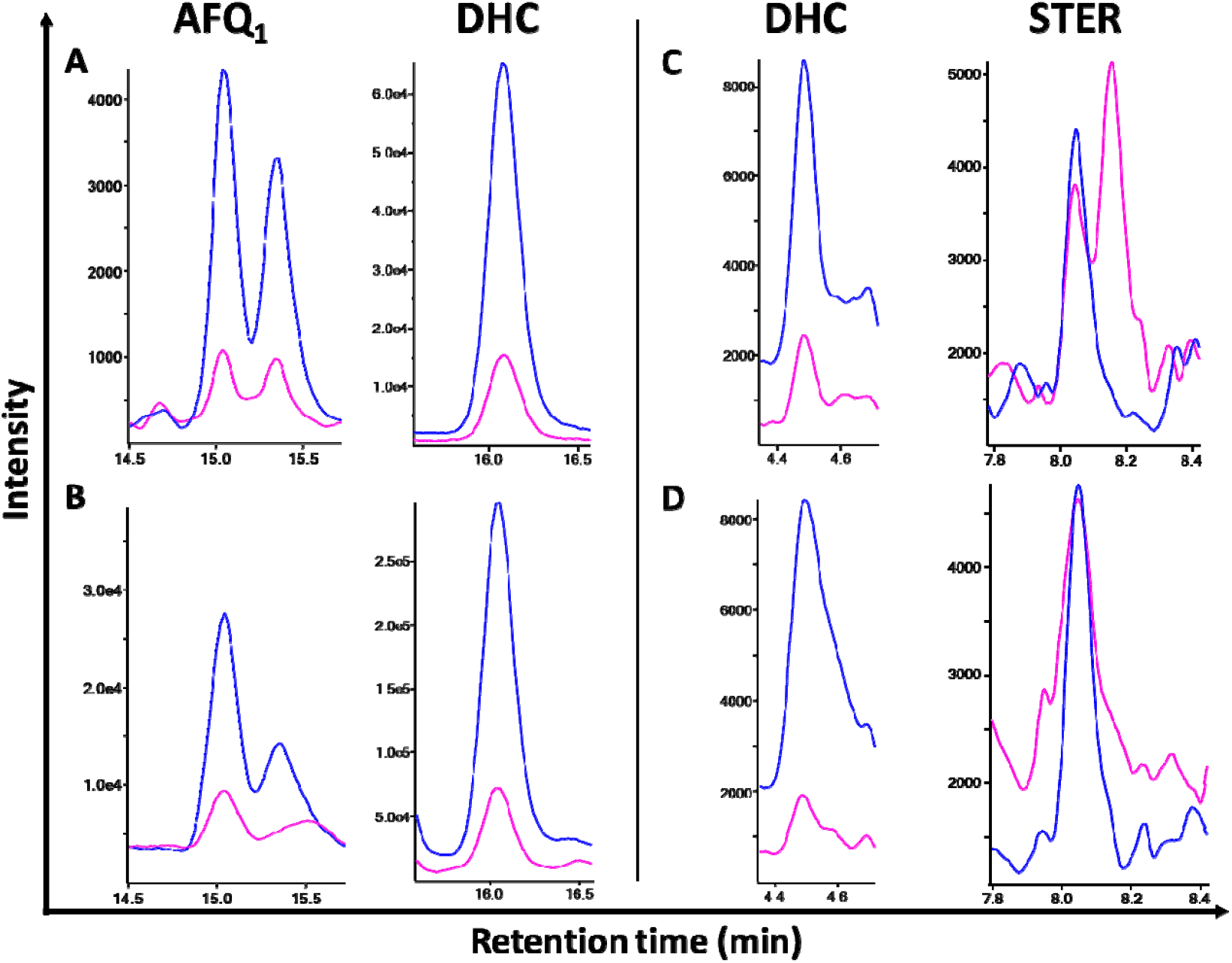
MRM-chromatograms of a matrix-matched standard in urine (A), and an infant urine sample (B) of aflatoxin Q_1_ (AFQ_1_) and dihydrocitrinone (DHC), respectively. In addition, MRM-chromatograms of a matrix-matched standard in breast milk (C) and a breast milk sample (D) of DHC and sterigmatocystin (STER), respectively, are shown. Quantifier, qualifier and retention time are given for each mycotoxin: AFQ_1_ (*m/z* 329.0–282.9; *m/z* 329.0–175.0; RT: 15.1 min) and DHC (*m/z* 265.0–221.1; *m/z* 265.0–246.9; RT: 16.1 min) in urine; DHC (*m/z* 265.0–177.0; *m/z* 265.0–203.0; RT: 4.5 min) and STER (*m/z* 325.1–281.1; *m/z* 325.1–310.2; RT: 8.1 min) in breast milk.

### 3.3 Distribution of mycotoxins in plate-ready complementary food

Sixty-five fungal metabolites, of which 14 were mycotoxins (Table 3), and 25 other secondary metabolites (Table S1) were quantified in the samples of plate-ready complementary food. Aflatoxin B_1_, B_2_ and G_1_ were detected, but not AFG_2_. Total AFs contaminated 21% of food samples of which the mean was 6.0 µg/kg (range: 1.0–16.2 µg/kg). The mean level of AFG_1_ (12 µg/kg) deriving from two *ogi* samples was three-fold higher than that of AFB_1_ (3 µg/kg). The incidence of FUM (sum of fumonisins B series (B_1_ and B_2_) was 29%, of which the mean was: 48.8 µg/kg (range 7.9–194 µg/kg). BEA (incidence: 79%; mean: 3.8 µg/kg) was the most frequently detected mycotoxin in the food samples. Enniatins, ZEN, MON and DON also occurred in 19, 10, 7 and 5% of the foods, respectively. With respect to occurrence of the toxins in food types, the highest mean concentrations of BEA (19.4 µg/kg), enniatins (9.6 µg/kg) and FUM (63.1 µg/kg), were found in infant cereal. Of the three food types, *ogi* contained the highest mean levels of AF (6.5 µg/kg) and DON (61.5 µg/kg). AF and MON were not found in infant cereal. *Ogi* and *tombran* contained more diverse mycotoxins than infant cereal (Table 3).

**Table 3.** Mycotoxins in plate-ready complementary food fed to non-exclusively breastfed infants in Ogun state, Nigeria.

| Mycotoxins | LOD (µg/kg) | LOQ (µg/kg) | All samples (n=42) |  |  | Infant cereal (n=6) |  | Ogi (n=26) |  | Tombran (n=10) |  |
| --- | --- | --- | --- | --- | --- | --- | --- | --- | --- | --- | --- |
|  |  |  | N (%)<sup>a</sup> | Range<sup>b</sup> | Mean<sup>b</sup>±SD | N | Mean<sup>b</sup> | N | Mean<sup>b</sup> | N | Mean<sup>b</sup> |
| Aflatoxin B<sub>1</sub> | 0.24 | 0.72 | 9 (21.4) | 1.0–7.0 | 3.0 ± 2.0 | 0 | <LOD | 6 | 2.5 | 3 | 4.1 |
| Aflatoxin B<sub>2</sub> | 0.4 | 1.2 | 1 (2.4) | 1.2 | 1.2 | 0 | <LOD | 0 | <LOD | 1 | 1.2 |
| Aflatoxin G<sub>1</sub> | 0.32 | 0.96 | 2 (4.8) | 12.0–12.1 | 12.1 ± 0.0 | 0 | <LOD | 2 | 12.1 | 0 | <LOD |
| **Total aflatoxin<sup>c</sup>** | - | - | **9 (21.4)** | **1.0–16.2** | **5.8 ± 6.3** | **0** | **<LOD** | **6** | **6.5** | **3** | **4.5** |
| Beauvericin | 0.008 | 0.024 | 33 (78.6) | 0.1–116 | 3.8 ± 20.1 | 6 | 19.4 | 19 | 0.3 | 8 | 0.3 |
| Deoxynivalenol | 1.2 | 3.6 | 2 (4.8) | 52.9–61.5 | 57.2 ± 6.1 | 1 | 52.9 | 1 | 61.5 | 0 | <LOD |
| Enniatin A<sub>1</sub> | 0.032 | 0.096 | 7 (16.7) | 0.1–5.7 | 1.3 ± 2.0 | 2 | 3.3 | 1 | 0.9 | 4 | 0.4 |
| Enniatin B | 0.024 | 0.072 | 8 (19.0) | 0.03–6.9 | 2.1 ± 3.0 | 2 | 6.9 | 1 | 1.1 | 5 | 0.4 |
| Enniatin B<sub>1</sub> | 0.04 | 0.12 | 6 (14.3) | 0.4–14.3 | 4.1 ± 5.3 | 2 | 9.6 | 1 | 2.2 | 3 | 1.1 |
| Enniatin B<sub>2</sub> | 0.04 | 0.12 | 3 (7.1) | 0.09–0.34 | 0.22 ± 0.13 | 2 | 0.29 | 0 | <LOD | 1 | 0.09 |
| Fumonisin A<sub>1</sub> | 2 | 6.0 | 1 (2.4) | 2.0 | 2.0 | 0 | <LOD | 1 | 2.0 | 0 | <LOD |
| Fumonisin B<sub>1</sub> | 3.2 | 9.6 | 11 (26.2) | 13.8–114 | 35.6 ± 32.2 | 2 | 54.2 | 7 | 35.3 | 2 | 18.0 |
| Fumonisin B<sub>2</sub> | 2.4 | 7.2 | 8 (19.0) | 7.9–80.3 | 24.3 ± 24.0 | 1 | 17.8 | 6 | 26.7 | 1 | 15.8 |
| **Total fumonisin<sup>d</sup>** | - | - | **12 (28.6)** | **7.9–194** | **48.8 ± 52.9** | **2** | **63.1** | **8** | **50.9** | **2** | **25.9** |
| Moniliformin | 1.6 | 4.8 | 3 (7.1) | 7.2–10.6 | 8.6 ± 1.8 | 0 | <LOD | 0 | <LOD | 3 | 8.6 |
| Zearalenone | 0.12 | 0.36 | 4 (9.5) | 0.5–6.8 | 2.4 ± 3.0 | 1 | 0.6 | 2 | 1.1 | 1 | 6.8 |

<sup>a</sup>Number (and percentage) of samples containing the mycotoxin. <sup>b</sup>Mean (and standard deviation) of mycotoxin concentrations expressed in µg/kg.
<sup>c</sup>Sum of aflatoxins B<sub>1</sub>, B<sub>2</sub> and G<sub>1</sub>.
<sup>d</sup>Sum of fumonisins B<sub>1</sub> and B<sub>2</sub>.

**Table 4.**
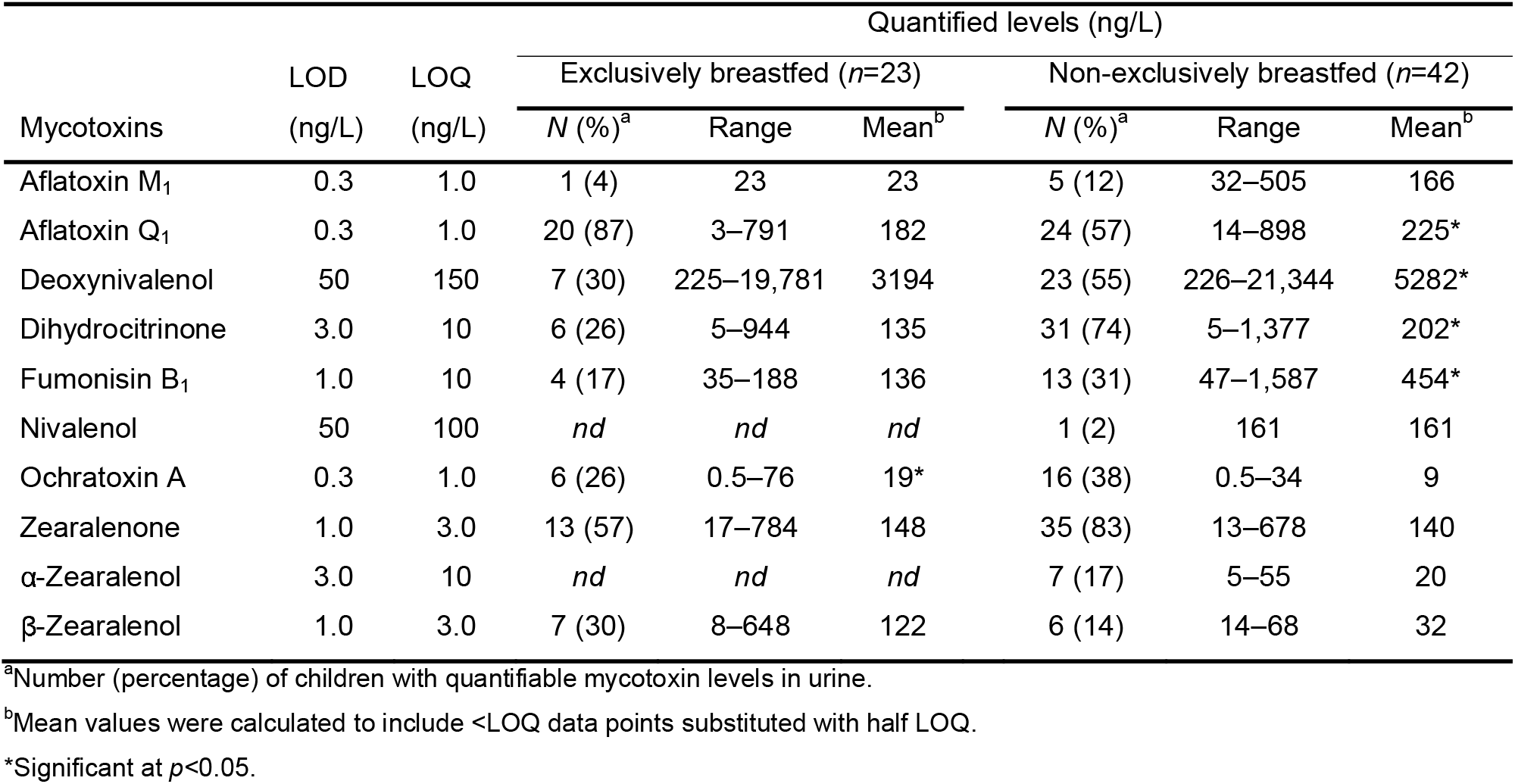
Mycotoxins in urines of exclusively breastfed (EB) and non-exclusively breastfed (NEB) infants in Ogun state, Nigeria.

| Mycotoxins | LOD<br>(ng/L) | LOQ<br>(ng/L) | Quantified levels (ng/L) |  |  |  |  |  |
| --- | --- | --- | --- | --- | --- | --- | --- | --- |
|  |  |  | Exclusively breastfed (*n*=23) |  |  | Non-exclusively breastfed (*n*=42) |  |  |
|  |  |  | *N* (%)<sup>a</sup> | Range | Mean<sup>b</sup> | *N* (%)<sup>a</sup> | Range | Mean<sup>b</sup> |
| Aflatoxin M<sub>1</sub> | 0.3 | 1.0 | 1 (4) | 23 | 23 | 5 (12) | 32–505 | 166 |
| Aflatoxin Q<sub>1</sub> | 0.3 | 1.0 | 20 (87) | 3–791 | 182 | 24 (57) | 14–898 | 225* |
| Deoxynivalenol | 50 | 150 | 7 (30) | 225–19,781 | 3194 | 23 (55) | 226–21,344 | 5282* |
| Dihydrocitrinone | 3.0 | 10 | 6 (26) | 5–944 | 135 | 31 (74) | 5–1,377 | 202* |
| Fumonisin B<sub>1</sub> | 1.0 | 10 | 4 (17) | 35–188 | 136 | 13 (31) | 47–1,587 | 454* |
| Nivalenol | 50 | 100 | *nd* | *nd* | *nd* | 1 (2) | 161 | 161 |
| Ochratoxin A | 0.3 | 1.0 | 6 (26) | 0.5–76 | 19* | 16 (38) | 0.5–34 | 9 |
| Zearalenone | 1.0 | 3.0 | 13 (57) | 17–784 | 148 | 35 (83) | 13–678 | 140 |
| α-Zearalenol | 3.0 | 10 | *nd* | *nd* | *nd* | 7 (17) | 5–55 | 20 |
| β-Zearalenol | 1.0 | 3.0 | 7 (30) | 8–648 | 122 | 6 (14) | 14–68 | 32 |

<sup>a</sup>Number (percentage) of children with quantifiable mycotoxin levels in urine.
<sup>b</sup>Mean values were calculated to include <LOQ data points substituted with half LOQ.
\*Significant at *p*<0.05.

### 3.4 Mycotoxins in urines of infants

#### 3.4.1 Overview of the distribution of mycotoxins in infant urines

At least one mycotoxin was detected in 64/65 (99%) of the urine samples, with 10 distinct mycotoxins observed, including aflatoxins (AFM_1_ and AFQ_1_), fumonisin B_1_, DON, DHC, NIV, OTA, zearalenones (ZEN, *α*-ZEL, and *β*-ZEL), representing seven classes of mycotoxin (Table 4). The detected aflatoxins were two metabolites: AFM_1_ (6/65, 9% of samples; mean of detected (MoD): 140 ng/L; range: 23–505 ng/L) and AFQ_1_ (44/65, 68% of samples; MoD: 210 ng/L; range: 3–898 ng/L). Two infants had only AFM_1_ detected, with no AFQ_1_, thus in total 46/65 (71%) of the infants had been exposed to dietary aflatoxin. Only one of the fumonisin class of mycotoxins was detected, FB_1_ (17/65, 26% of samples; MoD: 380 ng/L; range: 35–1587 ng/L). OTA, ZEN, DON and DHC were also frequently detected (Table 4). The chromatograms of AFQ_1_ and DHC detected in urine are given in Figure 1(A–B). Mycotoxins in urine were higher in females versus males, though no statistically significant differences in frequency or concentration for individual toxins was observed (data not shown).

#### 3.4.2 Mycotoxins in exclusively breastfed (EB) and non-exclusively breastfed (NEB) infants’ urine

Eight of the 10 mycotoxins were observed frequently in NEB infants compared to EB infants: AFM_1_ (12 vs 4%), DON (55 vs 30%), DHC (74 vs 26%), FB_1_ (31 vs 17%), OTA (38 vs 26%), ZEN (83 vs 57%), respectively, and NIV and α-ZEL which were only detected in NEB infants’ urine (Table 4). The recorded detection frequencies of the mycotoxins resulted in a 46– 300% increase from EB to NEB cohorts (Figure 2). There were however reductions in the frequencies of detection for AFQ_1_ (NEB: 53 vs EB: 87%) and *β*-ZEL (NEB: 14 vs EB: 30%). The mean concentrations in urine with detectable mycotoxins were higher, for seven of the 10 mycotoxins, in NEB versus EB infants: AFM_1_ (166 vs 23 ng/L), AFQ_1_ (225 vs 182 ng/L), DON (5,282 vs 3,194 ng/L), DHC (202 vs 135 ng/L), FB_1_ (454 vs 136 ng/L), NIV (161 ng/L vs non-detect) and α-ZEL (20 ng/L vs non-detect), respectively, see Table 4; resulting in 124–721% increase (Figure 2). However, significant (*p*<0.05) higher mean levels were only recorded for AFQ_1_, DON, DHC and FB_1_ in favour of NEB infants, and for OTA in favour of EB infants (Table 4).

**Figure 2.**
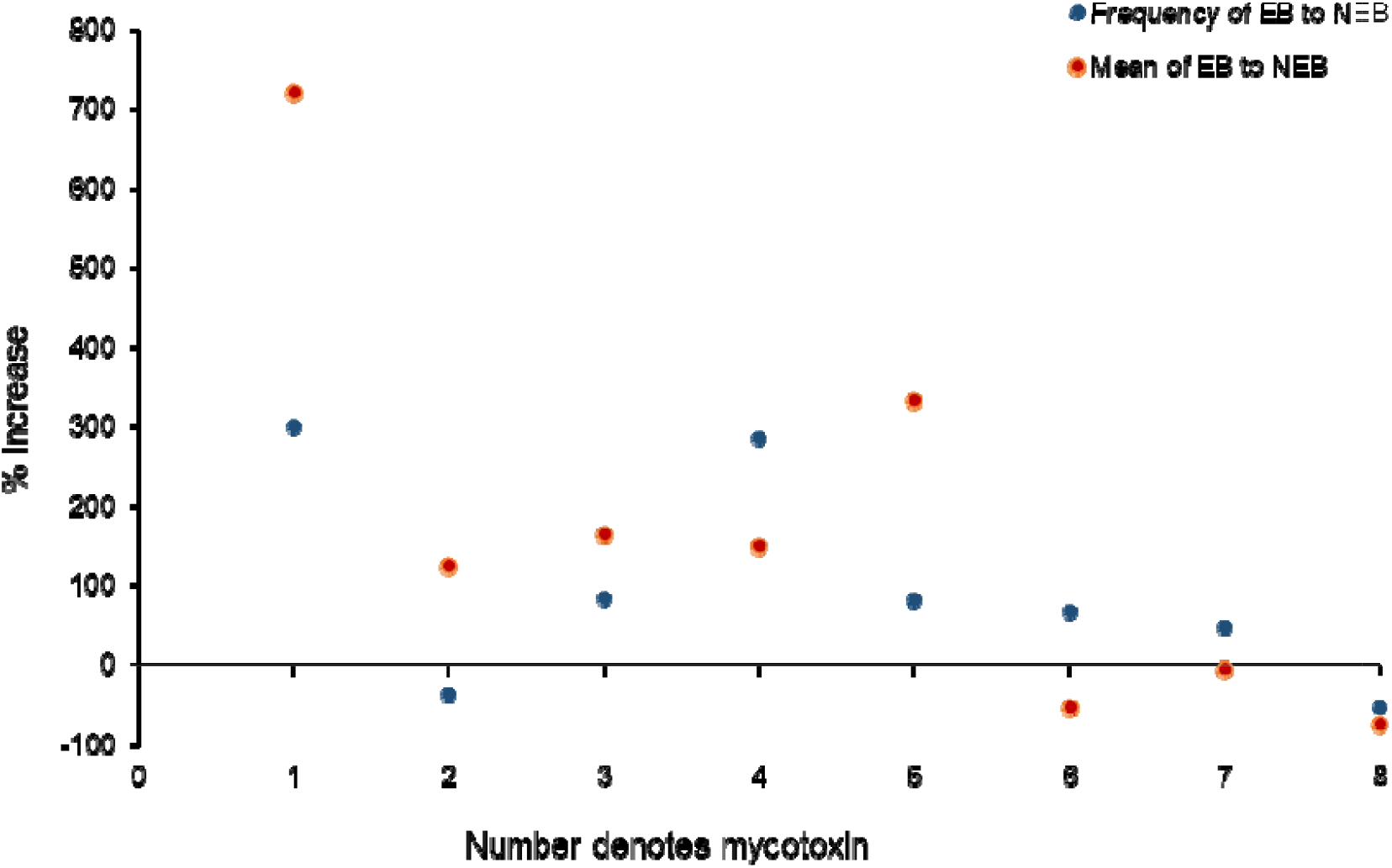
Scatter plot for percentage increases in detection frequencies and means of detects of mycotoxins in urine of non-exclusively breastfed (NEB) vs. exclusively breastfed (EB) infants in Ogun state, Nigeria. Numerals on x-axis denote mycotoxins: 1 = aflatoxin M_1_; 2 = aflatoxin Q_1_; 3 = deoxynivalenol; 4 = dihydrocitrinone; 5 = fumonisin B_1_; 6 = ochratoxin A; 7 = zearalenone; 8 = β-zearalenol.

### 3.5 Mycotoxin co-occurrence and co-exposures

The numbers of different classes of mycotoxins detected in each of breast milk, complementary food and urine were determined (Table 5). All breast milk samples contained more than one and up to five mycotoxin classes; 41% of samples had four or more different mycotoxins detected, though overall mycotoxin concentrations were low. The primary combinations in the mycotoxin mixtures in breast milk included AME, BEA, enniatins and OTA. Aflatoxins were observed as part of 4/22 (18%) mixtures of toxins. Fumonisins in breast milk were not measured in the present study.

**Table 5.**
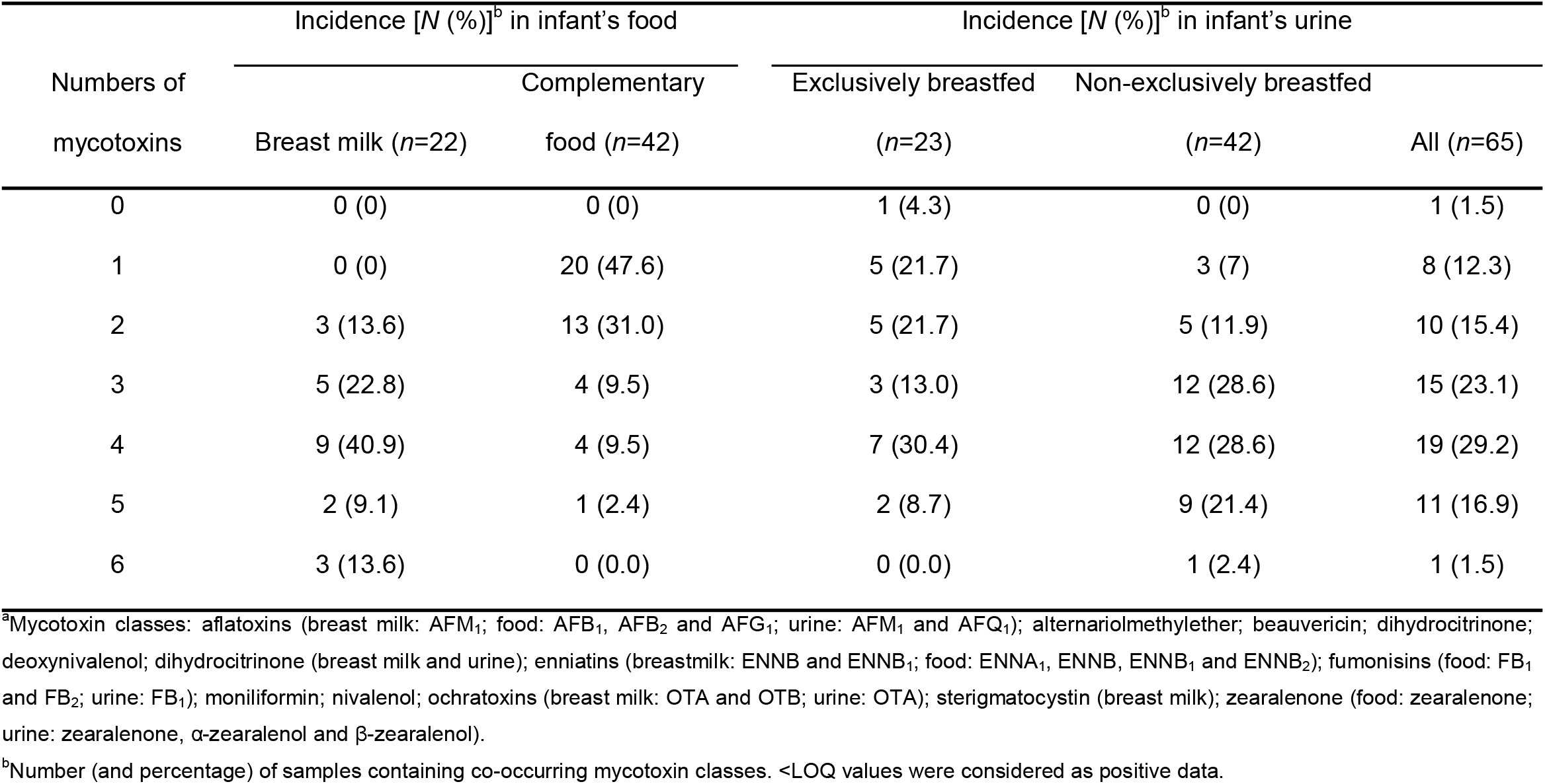
Co-occurrence of mycotoxin classes^a^ in breast milk, complementary food and urine.

Mycotoxin mixtures, of 2–5 toxins, were observed in just over half (22/42; 52%) of the plate-ready complementary food samples. The observed mixtures in the complementary food samples predominantly involved three classes of mycotoxins: BEA in 12 mixtures, FUM in nine mixtures and AF in eight mixtures. Of these mixtures, five samples with three or more toxins contained both AF and FUM.

Mixtures of 2–6 mycotoxins were observed in about 86% of the urines, with NEB typically having more urine samples with mixtures (93%, with 2–6 toxins) compared to EB infants (74% with 2–5 toxins). The most frequently occurring classes of toxins in mixtures were ZEN (45/65; 69%), AF (42/65; 65%), DHC (41/65; 63%) and DON (30/65; 46%). AF and FUM in the same urine occurred in 13/65 (20%). One infant from the NEB cohort was co-exposed to seven mycotoxins and metabolites belonging to six of the assessed mycotoxin classes except NIV (Figure 3).

**Figure 3.**
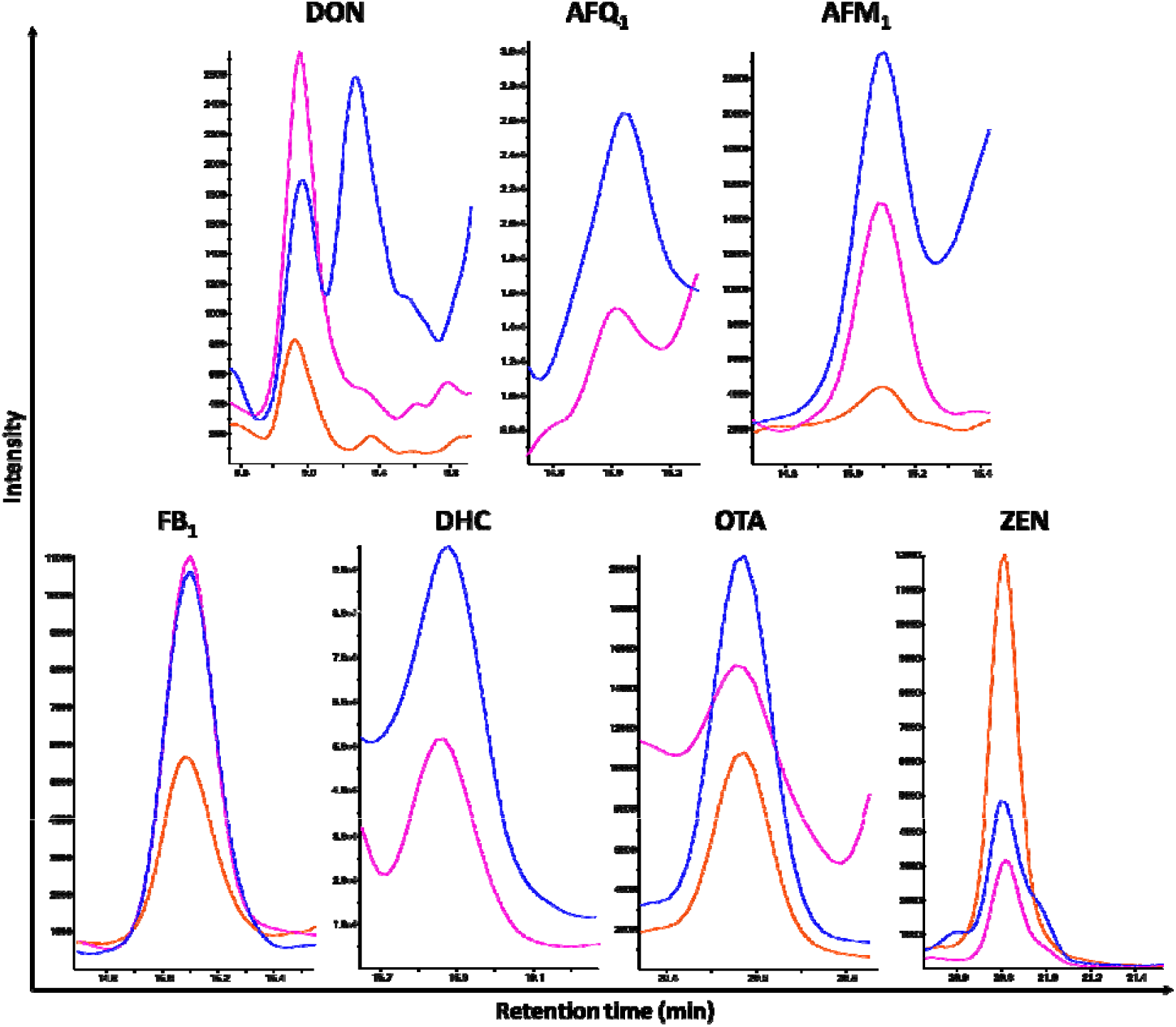
MRM-chromatogram of one non-exclusively breastfed infant urine sample from Ogun state, Nigeria, containing a mixture of seven mycotoxins belonging to six mycotoxin classes. Quantifier, qualifier, internal standard (IS) and retention time are given for each mycotoxin: DON (*m/z* 355.1–246.9; *m/z* 355.1–137.9; *m/z* 370.1–278.8; RT: 9.0 min), AFQ_1_ (*m/z* 329.0–282.9; *m/z* 329.0–175.0; no IS; RT: 15.0 min), AFM_1_ (*m/z* 329.1–273.2; *m/z* 329.1–229.1; *m/z* 346.0– 288.2; RT: 15.1 min), FB_1_ (*m/z* 722.5–334.4; *m/z* 722.5–352.3; *m/z* 756.3–356.3; RT: 15.1 min), DHC (*m/z* 265.0–221.1; *m/z* 265.0–246.9; no IS; RT: 15.9 min), OTA (*m/z* 404.0–239.0; *m/z* 404.0–358.0; *m/z* 424.2–250.1; RT: 20.6 min), and ZEN (*m/z* 317.1–175.0; *m/z* 317.1–131.0; *m/z* 335.2–185.1; RT: 20.8 min).

Paired frequencies of mycotoxin detected in the matrices from each cohort were compared. In the EB cohort, contamination frequencies in breast milk and urine were AFM_1_/AFM_1_ (18%/4%), AFM_1_/AFQ_1_ (18%/87%), DHC (27%/26%) and OTA (64%/26%). The contamination frequencies in complementary food vs urine in the NEB cohort were AFB_1_/AFM_1_ (27%/12%), AFB_1_/AFQ_1_ (27%/57%), DON (5%/55%), FB_1_ (29%/31%) and ZEN (10%/83%).

## 4. Discussion

Infants’ transition from breastfeeding to complementary food is an essential nutritional requirement. However, it is important to understand if this transition creates opportunities for differential exposure to diverse classes of fungal toxins and also other food and environmental contaminants (i.e., the exposome). The diversity of mycotoxins in breast milk is still in the exploratory stage, though several mycotoxins have been observed, most notably aflatoxins; however, food measurements are by far more established to date despite several limitations (IARC, 2012a, 2012b, 2015). This study provides a measure of multiple mycotoxins in both breast milk and plate-ready complementary foods, and then compares patterns of these mycotoxins in urine from infants that are either EB or partially receiving breast milk and complementary foods, NEB.

In this study, the analytical sensitivity for measuring mycotoxins in breast milk was extremely high (low LOD), and thus the frequency of toxin detection was quite high, providing unique insights into chronic background level exposure patterns. All 22 samples had two or more toxins, though overall the concentrations would be regarded as low. The lipophilic aflatoxins such as AFB_1_ or AFG_1_ are commonly detected in grains in sub-Saharan Africa (Ezekiel et al., 2018a), but were not observed in breastmilk. The AFB_1_ metabolite, AFM_1_, was observed in four samples and at a level below the LOQ (4 ng/L). This data suggests that breast milk is not providing significant exposure to the most carcinogenic of the mycotoxins for participants of this study. AFM_1_ in breast milk occurred at lower mean concentrations than those previously reported from Sierra Leone (800 ng/L, Jonsyn et al., 1995), Egypt (60 ng/L, Polychronaki et al., 2007), Nigeria (35 ng/L, Adejumo et al., 2013) and Ecuador (45 ng/L, Ortiz et al., 2018), but similar to levels reported in two samples from Brazil (0.3 and 0.8 ng/L, Iha et al., 2014).

Prior to the present study, there were no reports on the occurrence of CIT and its metabolites as well as STER in human breast milk (Ali and Degen, 2019; Cherkani-Hassani et al., 2016; Warth et al., 2016). Thus, to the best of our knowledge, we present the first report of DHC and STER occurrences in human breast milk. DHC, mostly considered a detoxification product of CIT but also co-detected with CIT in food samples (Ezekiel et al., 2020; Ojuri et al., 2018, 2019), is indicative of CIT exposure (Ali and Degen, 2019; Follmann et al., 2014). The concentrations of DHC in breast milk samples analyzed in the present study were low, and as much as 10 times lower than the concentrations found in urine. However, the presence of DHC in both biological fluids compel us to hypothesize that CIT intake is common in Nigeria (Akinfala et al., 2020; Ezekiel et al., 2020; Ojuri et al., 2018, 2019). Furthermore, it agrees with the suggestions of Ali and Degen (2019) who retrospectively estimated high CIT exposure assessment/intake above the “level of no concern for nephrotoxicity” of 0.2 μg/kg bwt/day set by EFSA (EC, 2014; EFSA, 2012) in participants in a previous urinary biomarker study in Nigeria (Šarkanj et al., 2018). Due to sparse data on its occurrence and toxicity, there is no regulation on maximum levels (ML) of STER in food, including infant diets, in most countries, Nigeria inclusive (EC, 2006; Oplatowska-Stachowiak et al., 2018). Nonetheless, this mycotoxin may be carcinogenic in humans (EFSA, 2013; IARC, 1987), as such, its detection in breast milk samples provides a basis for the consideration of more investigations into the occurrence and regulation of this mycotoxin in foods intended for infants and young children.

The mean level of the nephrotoxic OTA in the analyzed breast milk was 50 times lower than the maximum tolerable limit of 500 ng/L set for infant food in Europe (EC, 2006). Additionally, the recorded level was four to five times lower than those reported in Chile (mean: 44 ng/L, Munoz et al., 2014) and Bolivia (mean: 53 ng/L, Ferrufino-Guardia et al., 2019) but nearly two times higher than the mean level reported in Italy (mean: 6.0 ng/L, Turconi et al., 2004). The identification of BEA and EnnB in human breast milk agrees with previous reports from Spain (Rubert et al., 2014) and Nigeria (Braun et al., 2018). The mean concentration of AME in the present study is, however, lower than the mean level (11.5 ng/L) in a previous study from Nigeria (Braun et al., 2020). Due to poor analytical recoveries at the outset, fumonisins were not included in the breast milk analysis. The observed variations in mycotoxin levels in breast milk from our study compared to previous reports may be due to any or several of these factors: differences in study population (geography and size), dietary patterns, metabolic status of mothers, lactation stage, or even the uncertainty related to comparing data from distinct analytical methods.

Mycotoxins have been previously reported in complementary foods (Cappozzo et al., 2017; Chuisseu Njamen et al., 2018; Juan et al., 2014; Kamala et al., 2016; Kolakowski et al., 2016; Ojuri et al., 2018, 2019; Oueslati et al., 2018). The detection of more diverse spectra of mycotoxins (AF, BEA, EnnB, FUM, MON and ZEN) in the traditionally processed complementary foods (*ogi* and *tombran*) compared to the industrially processed infant cereal agrees with an earlier report from our group on mycotoxins in complementary foods from Nigeria (Ojuri et al., 2018, 2019). AFs were detected in about 20% of the food samples, but notably not in the infant cereal. The mean of these samples with detectable AFs in traditionally processed complementary foods exceeded the 4 µg/kg maximum level for total AF in Nigerian baby food (Ojuri et al., 2018). The presence of AF in locally processed complementary foods but not in commercial infant cereal is in keeping with the report from Cameroon (Chuisseu Njamen et al., 2018). However, the levels found in the present study were lower than those previously reported in locally processed baby foods from Cameroon (mean: 177 ug/kg, Chuisseu Njamen et al., 2018) and Nigeria (mean: 104 µg/kg, Ojuri et al., 2018), but similar to Ghana (mean: 8.3 µg/kg, Blankson et al., 2019). Unlike most of the aforementioned reports, herein we analyzed plate-ready food for a more precise food contamination data because it considers food ingested after final preparatory steps such as dilution and cooking (Chuisseu Njamen et al., 2018; Ezekiel et al., 2019). Clearly, the methods employed at household levels, often characterized by poor food processing practices, influenced the mycotoxin contents in the traditionally processed complementary foods. The fact that we could not detect AF in the infant cereal could be attributed to tailoring of production by industrial processors to meet regulatory standards for AF.

FUM was detected in all the complementary food types. However, the mean concentrations we found were lower than the European Union ML of 200 µg/kg for baby food (EC, 2006) and the mean level (2808 µg/kg) reported in Tanzania (Kamala et al., 2016). FUM is not regulated in baby foods in Nigeria. Nevertheless, the observed co-occurrence of FUM with AF in about 20% of the complementary foods is of concern, because these two mycotoxins are suggested to negatively influence linear growth in children (IARC, 2015; Shirima et al., 2015). Meanwhile, a potential role of additional toxins in mixtures is unknown.

Several mycotoxins were detected in the children’s urine similar to previous reports from Cameroon (Ediage et al., 2013), Nigeria (Ezekiel et al., 2014), Spain (Rodriguez-Carrasco et al., 2014) and Belgium (Heyndrickx et al., 2015) where the simultaneous exposure to more than two mycotoxins were detected. The incidences and analytical concentrations of commonly found mycotoxins in children urine were compared with data from previous studies. Overall, ZEN (incidence: 74%), a potent estrogenic mycotoxin (IARC 2012b, 2015), was the most frequently detected mycotoxin in the urine samples. Two ZEN metabolites, α-ZEL and β-ZEL, were detected in the urine samples albeit at lower frequencies (11% and 20%, respectively) compared to the parent compound. This is in contrast to the reports of Heyndrickx et al. (2015) wherein ZEN and its metabolites were not detected in children urine from Belgium. The incidence of ZEN in the present study was, however, higher than the incidence (4%, 9/220) previously reported in Cameroonian children employing a method of lower sensitivity, whereas the mean level was higher in the Cameroonian children (970 ng/L) (Ediage et al., 2013) than in the present study (142 ng/L). Similarly, the incidence of DHC in the present study (57%, 37/65) was higher, although with a lower mean concentration of 212 ng/L, than the 14% (20/142) incidence and mean of 490 ng/L reported in urine of adults and children in Haiti (Gerding et al., 2015). Additionally, the incidence of DHC in the present study was much higher than the 6% (7/124) incidence reported for DHC in urine of Belgian children (Heyndrickx et al., 2015). The parent toxin CIT was not detected in any urine sample, which agrees with the reports from Haiti but negates the Belgian study that found both parent compound (CIT) and its metabolite (DHC). Regardless, the higher incidence of DHC recorded in the present study further suggests that CIT exposure is widespread in Nigerian children. DON (46%, 30/65) occurrence in urine in the present study was lower than those previously reported in adults from France (99%, 75/76, Turner et al., 2010), pregnant women (100%, 85/85, Hepworth et al., 2012) and children in the United Kingdom (100%, 40/40, Papageorgiou et al., 2018; 100%, 21/21, Gratz et al., 2020), but more similar to in the reports from Egypt (68%, 63/93, Piekkola et al., 2012) and Iran (72%, 79/110, Turner et al., 2012b). Infant data on DON in urine are mostly lacking. In the present study, urinary OTA (34%, 22/65; mean: 14 ng/L) was lower than the previous report in Portuguese children (93%, 79/85; mean: 20 ng/L) (Silva et al., 2019). Similarly, urinary FB_1_ incidence of 26% (17/65) recorded in the present study was lower than the 96% incidence reported in a study comprising 147 Tanzanian children’s urine (Shirima et al., 2013) but was higher than the incidence (2%, 1/50) reported in Nepalese children (Mitchell et al., 2016).

To date, AFM_1_ has been the most reported urinary AF metabolite and biomarker in children (Ayelign et al., 2017; Chen et al., 2017; Ediage et al., 2013; Ezekiel et al., 2014; Gerding et al., 2015; Polychronaki et al., 2008; Sanchez and Diaz, 2019; Schwartzbord et al., 2016). However, in the present study, AFQ_1_ was found to be the major aflatoxin metabolite (both in frequency and concentration), occurring in 44/65 (68%) of the children’s urine samples, whereas AFM_1_ was detected in 6/65 (9%) of urine samples, four of which had both AFM_1_ and AFQ_1_. Previous reports on AFQ_1_ occurrence in human urine were on adolescents and adults 15–64 years of age in China and The Gambia (Groopman et al., 1992a, 1992b; Mykkanen et al., 2005; Wang et al., 2001). Consequently, to the best of our knowledge, AFQ_1_ is reported for the first time in the urine of children in any setting. Urinary AFM_1_ and AF-N7-guanine are established biomarkers for aflatoxin exposure; however, these were established in adults (Groopman et al., 1992a). Given the role of CYP3Aa4 in AFQ_1_ formation (Wild and Turner, 2002) and the known significant ontogeny in xenobiotic metabolizing enzymes (Blake et al., 2005), involved in AFB_1_ biotransformation, it would be useful to better understand the relationship between dose and the various metabolites of aflatoxins in urine of infants. Nevertheless, the recorded higher incidence of AFQ_1_ in the urine samples compared with AFM_1_ informs us that aflatoxin exposure in young children may be underreported.

Comparing the two cohorts of infants, higher mean levels of seven out of the 10 detected mycotoxins were found in the NEB urine compared to the EB urine, with statistically significant levels recorded for four of these mycotoxins including AFQ_1_, DON, DHC and FB_1_. Notably, increases in the mean mycotoxin levels in urine moving from EB regime to the NEB regime reached 721 and 334% for AFM_1_ and FB_1_, respectively. Additionally, the NEB cohort generally recorded higher diversities of mycotoxin mixtures in urine compared to the EB cohort. Here, we show that transition from EB to NEB, involving a change in diet, modulates contaminant exposure resulting in a large variation of multiple mycotoxin exposures in infants. This primarily owes to the higher intake of complementary foods, which are usually made from mycotoxin-prone cereals, nuts and oil seeds (Chuisseu Njamen et al., 2018; Juan et al., 2014; Kamala et al., 2016; Kimanya et al., 2009; 2010, 2014; Ojuri et al., 2018, 2019; Okeke et al., 2015). These observations are strongly suggestive that exclusive breastfeeding provides a relatively, but not completely, protected period from mycotoxin exposure.

The spectra and combinations of mycotoxins detected in the three specimens examined in this study further suggest frequent exposure in Nigerian children, especially in the NEB category. Obviously, urine contained higher number of mycotoxins and their mixtures compared to complementary foods and breast milk. Comparison of the paired contamination frequencies in both cohorts, especially in the NEB, favored urine with higher frequencies detected for aflatoxins, FB_1_ and ZEN compared to complementary foods. Further, some mycotoxins occurred only in the biological fluids reiterating the importance of preferring biomonitoring approaches over food contamination data for exposure assessment in humans (Ali and Degen, 2019; Turner et al., 2012; Vidal et al., 2018; Warth et al., 2013). A typical example is DHC and OTA that were detected only in the biological fluids (breast milk for the EB group and urine from both cohorts). Although CIT/DHC was not detected in the examined complementary foods in the present study, it was found in 74% of the urines of EB children. We did not sample milk for the NEB cohort due to reluctance in provision of milk by mothers in this group; nevertheless, it is possible that the DHC levels in the urines of both cohorts originated from the lactating mothers’ milk since this metabolite or its parent toxin was not found in the complementary food. CIT levels in grains (especially maize) and complementary food in Nigeria have recently been shown to be high reaching 16,000 µg/kg (Ezekiel et al., 2020; Ojuri et al., 2018, 2019; Okeke et al., 2015, 2018). Fermented cocoa beans in Nigeria, which could also be processed and used in the preparation of foods for mothers as well as for infants and young children, have also recently been reported to contain CIT (Akinfala et al., 2020). Put together, only aflatoxins, DHC and OTA were found in both breast milk and urine of the EB cohort, while aflatoxins, DON, FB_1_ and ZEN were detected in both complementary food and NEB children’s urine.

An additional important finding was the detection of BEA and EnnB in complementary food and breast milk but not in urine because the urinary analytical method did not include both mycotoxins. EnnB was recently detected in adult human urine from Italy (Rodriguez-Carrasco et al., 2018); however, there is limited data on urinary BEA and EnnB in children. Considering the widespread detection of BEA and EnnB in foods in Nigeria, albeit at low concentrations (Abdus-Salaam et al., 2015; Adetunji et al., 2014; Ezekiel et al., 2016; Ojuri et al., 2018; Oyedele et al., 2017), and the documented *in vitro* data suggesting their roles in decreasing immune response to infections (Ficheux et al., 2013) as well as their interactions to cause additive myelotoxicity (Ficheux et al., 2012), the inclusion of both mycotoxins in future urinary biomonitoring studies, especially for children, is recommended.

In this study, the levels of any given mycotoxin were typically modest compared to studies where associations with an individual mycotoxin and a given health outcome, e.g., stunting, was reported (IARC, 2015). However, the study reveals a clear transition in exposure from EB to NEB infants in terms of both level and frequency to mixtures. The overall health implication for children in the study region was not measured. Focusing on infant growth, *in vitro* studies using human intestinal cell lines revealed that DON selectively modulated intestinal transporters (Maresca et al., 2002) while FB_1_ decreased intestinal cell viability and proliferation (Minervini et al., 2014). Consequently, these mycotoxins may play interactive roles to affect nutrient absorption (Liew and Mohd-Redzwan, 2018) which may cumulatively contribute to AF induced growth faltering. Thus, future studies may consider exploring the type and extent of interactive effects the various recorded mixtures may have on this highly vulnerable population and their growth into adulthood.

## 5. Conclusion

This study provides a comprehensive account on and novel insights into multiple mycotoxin exposures in EB and NEB infants in a high-risk country, Nigeria, based on examination of three specimens (breast milk, complementary food and urine). One pitfall of this study is the non-collection of breast milk for the NEB cohort, leaving the conclusions on NEB cohort anchored more on the urine data, which served as a comparative matrix for both EB and NEB cohorts. However, data for breast milk from EB cohort suggested that no significant exposure to the most carcinogenic mycotoxins was recorded. Consequently, we report the presence of several uncommon mycotoxins in biological fluids (DHC and STER in breast milk; and AFQ_1_, being the dominant urinary aflatoxin metabolite). The higher incidence of AFQ_1_ in infant urine compared with AFM_1_ suggests that aflatoxin exposure in young children may be underreported. Second, the presence of up to six distinct mycotoxin classes are reported for the first time in breast milk and infant urine. Third, urines of NEB infants contained higher levels of mycotoxins and several mixtures of these toxins more frequently than in urines of EB infants. Thus, exposure to mycotoxins for infants less than 18 months of age is common in the region, with a significant transition in higher frequency and levels of exposure with the introduction of complementary foods. Consequently, it is recommended that mothers adhere strictly to measures with potential to reduce mycotoxin contamination in household diets; including maintaining compliance to WHO recommendations for exclusive breastfeeding for 6 months. Once foods are being introduced, adequate drying and proper storage of grains in airtight containers at household levels, sorting out of moldy, damaged and discolored grains prior to meal preparation, and diet diversification to include less mycotoxin prone food crops such as tubers are advised. In addition, prioritizing partnerships focused on driving interventions to mitigate exposures in vulnerable populations is important. Furthermore, attempts to investigate mycotoxin exposure and toxicity holistically in the context of the exposome and DOHaD paradigm should be encouraged.

## Data Availability

Data in this study are available on request.

## Acknowledgements

Authors are thankful to Chidinma Akwarandu and Linda Ogu for their assistance during sample collection, and Xiaomin Han, during urine sample preparation. Families who donated samples are deeply appreciated. The authors want to gratefully acknowledge the Mass Spectrometry Centre (MSC) of the Faculty of Chemistry at the University of Vienna and Sciex for providing mass spectrometric instrumentation to BW. This work was performed with the financial support of the University of Vienna, the City of Vienna Jubilee Funds (BOKU Research Funding, project MycoMarker) and the Austrian Science Fund (FWF): P 33188-B.

## Table captions

**Table S1.**
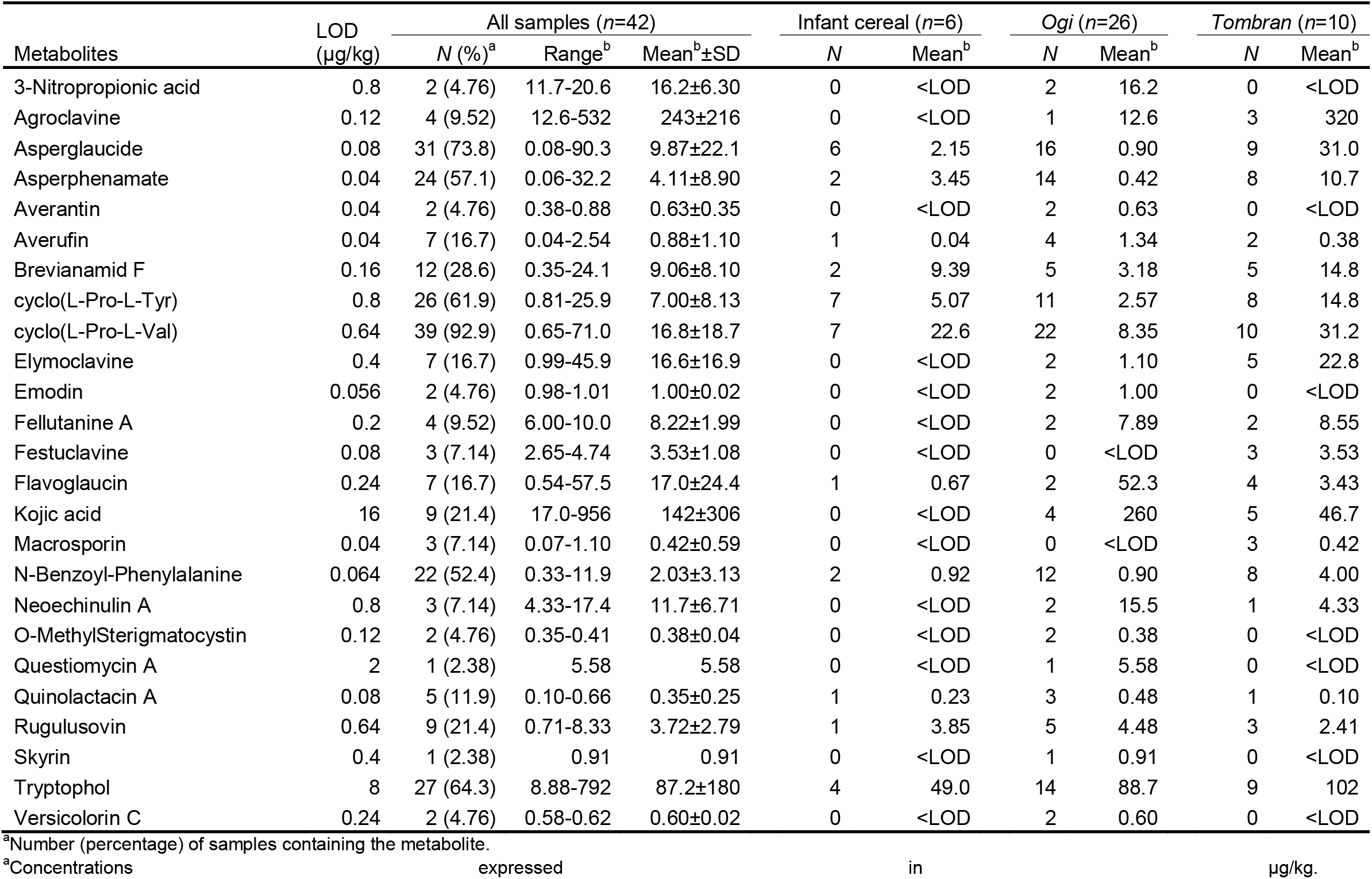
Fungal metabolites in plate-ready complementary food fed to non-exclusively breastfed infants in Ogun state, Nigeria.

| Metabolites | LOD<br>(µg/kg) | All samples (n=42) |  |  | Infant cereal (n=6) |  | Ogi (n=26) |  | Tombran (n=10) |  |
| --- | --- | --- | --- | --- | --- | --- | --- | --- | --- | --- |
|  |  | N (%)<sup>a</sup> | Range<sup>b</sup> | Mean<sup>b</sup>±SD | N | Mean<sup>b</sup> | N | Mean<sup>b</sup> | N | Mean<sup>b</sup> |
| 3-Nitropropionic acid | 0.8 | 2 (4.76) | 11.7-20.6 | 16.2±6.30 | 0 | <LOD | 2 | 16.2 | 0 | <LOD |
| Agroclavine | 0.12 | 4 (9.52) | 12.6-532 | 243±216 | 0 | <LOD | 1 | 12.6 | 3 | 320 |
| Asperglaucide | 0.08 | 31 (73.8) | 0.08-90.3 | 9.87±22.1 | 6 | 2.15 | 16 | 0.90 | 9 | 31.0 |
| Asperphenamate | 0.04 | 24 (57.1) | 0.06-32.2 | 4.11±8.90 | 2 | 3.45 | 14 | 0.42 | 8 | 10.7 |
| Averantin | 0.04 | 2 (4.76) | 0.38-0.88 | 0.63±0.35 | 0 | <LOD | 2 | 0.63 | 0 | <LOD |
| Averufin | 0.04 | 7 (16.7) | 0.04-2.54 | 0.88±1.10 | 1 | 0.04 | 4 | 1.34 | 2 | 0.38 |
| Brevianamid F | 0.16 | 12 (28.6) | 0.35-24.1 | 9.06±8.10 | 2 | 9.39 | 5 | 3.18 | 5 | 14.8 |
| cyclo(L-Pro-L-Tyr) | 0.8 | 26 (61.9) | 0.81-25.9 | 7.00±8.13 | 7 | 5.07 | 11 | 2.57 | 8 | 14.8 |
| cyclo(L-Pro-L-Val) | 0.64 | 39 (92.9) | 0.65-71.0 | 16.8±18.7 | 7 | 22.6 | 22 | 8.35 | 10 | 31.2 |
| Elymoclavine | 0.4 | 7 (16.7) | 0.99-45.9 | 16.6±16.9 | 0 | <LOD | 2 | 1.10 | 5 | 22.8 |
| Emodin | 0.056 | 2 (4.76) | 0.98-1.01 | 1.00±0.02 | 0 | <LOD | 2 | 1.00 | 0 | <LOD |
| Fellutanine A | 0.2 | 4 (9.52) | 6.00-10.0 | 8.22±1.99 | 0 | <LOD | 2 | 7.89 | 2 | 8.55 |
| Festoclavine | 0.08 | 3 (7.14) | 2.65-4.74 | 3.53±1.08 | 0 | <LOD | 0 | <LOD | 3 | 3.53 |
| Flavoglucin | 0.24 | 7 (16.7) | 0.54-57.5 | 17.0±24.4 | 1 | 0.67 | 2 | 52.3 | 4 | 3.43 |
| Kojic acid | 16 | 9 (21.4) | 17.0-956 | 142±306 | 0 | <LOD | 4 | 260 | 5 | 46.7 |
| Macrosporin | 0.04 | 3 (7.14) | 0.07-1.10 | 0.42±0.59 | 0 | <LOD | 0 | <LOD | 3 | 0.42 |
| N-Benzoyl-Phenylalanine | 0.064 | 22 (52.4) | 0.33-11.9 | 2.03±3.13 | 2 | 0.92 | 12 | 0.90 | 8 | 4.00 |
| Neoechinulin A | 0.8 | 3 (7.14) | 4.33-17.4 | 11.7±6.71 | 0 | <LOD | 2 | 15.5 | 1 | 4.33 |
| O-MethylSterigmatocystin | 0.12 | 2 (4.76) | 0.35-0.41 | 0.38±0.04 | 0 | <LOD | 2 | 0.38 | 0 | <LOD |
| Questiomyacin A | 2 | 1 (2.38) | 5.58 | 5.58 | 0 | <LOD | 1 | 5.58 | 0 | <LOD |
| Quinolactacin A | 0.08 | 5 (11.9) | 0.10-0.66 | 0.35±0.25 | 1 | 0.23 | 3 | 0.48 | 1 | 0.10 |
| Rugulusovin | 0.64 | 9 (21.4) | 0.71-8.33 | 3.72±2.79 | 1 | 3.85 | 5 | 4.48 | 3 | 2.41 |
| Skyrin | 0.4 | 1 (2.38) | 0.91 | 0.91 | 0 | <LOD | 1 | 0.91 | 0 | <LOD |
| Tryptophol | 8 | 27 (64.3) | 8.88-792 | 87.2±180 | 4 | 49.0 | 14 | 88.7 | 9 | 102 |
| Versicolorin C | 0.24 | 2 (4.76) | 0.58-0.62 | 0.60±0.02 | 0 | <LOD | 2 | 0.60 | 0 | <LOD |

<sup>a</sup>Number (percentage) of samples containing the metabolite.
<sup>a</sup>Concentrations expressed in µg/kg.

